# Adjuvant Immune Checkpoint Inhibitors After Radical Surgery for High-Risk Urothelial Carcinoma: *A Meta-Analysis*

**DOI:** 10.1101/2025.10.16.25338141

**Authors:** Stephen Adebayo Osasan, A. Alzahrani Hind, Ambrose Oche George, Mohammad Alshehri Jawaher, Nafie Khalid, Oluwafemi Ayodele Adefolalu, Abdulrahman Abdullah Alghamdi Shahd, Wisdom Ibrahim

## Abstract

**Background:** Patients with high-risk muscle-invasive urothelial carcinoma (MIUC) remain at substantial risk of recurrence following radical surgery. Adjuvant immune checkpoint inhibitors (ICIs) targeting the PD-1/PD-L1 axis have emerged as a potential strategy to reduce recurrence and improve survival outcomes.

**Methods:** We conducted a systematic review and meta-analysis of randomized controlled trials (RCTs) evaluating adjuvant ICIs versus observation or placebo in high-risk MIUC. Data were extracted for disease-free survival (DFS), overall survival (OS), distant metastasis-free survival (DMFS), recurrence-free survival beyond the urothelial tract (RFS-extraUT), and health-related quality of life (HRQoL). Standardized mean differences (SMDs) were used to express effect sizes with 95% confidence intervals (CIs), and pooled using inverse-variance weighting under common- and random-effects models. Heterogeneity and subgroup analyses were performed according to drug class, PD-L1 status, and clinical covariates.

**Results:** Nine study-level comparisons from four phase III RCTs (CheckMate 274, IMvigor010, AMBASSADOR, and related updates; n > 2,200) were included. Adjuvant ICIs significantly prolonged DFS compared with control (SMD –0.32; 95% CI –0.44 to –0.21; p < 0.001), with consistent benefits across subgroups and low heterogeneity (I² = 25%). OS benefit was emerging, with nivolumab demonstrating significant advantage at extended follow-up (HR 0.76; 95% CI 0.61–0.96), while pembrolizumab and atezolizumab yielded neutral OS results.

**Conclusions:** Adjuvant PD-1/PD-L1 inhibitors improve DFS and show emerging OS benefits without compromising HRQoL in high-risk MIUC. Nivolumab has demonstrated the most robust long-term survival evidence to date, supporting its role as standard of care. Further biomarker-driven trials are warranted to refine patient selection.

## 1.0 INTRODUCTION

Muscle-invasive urothelial carcinoma (MIUC) remains a highly aggressive malignancy with significant morbidity and mortality despite advances in surgical and perioperative management. Radical cystectomy or nephroureterectomy with lymph node dissection represents the cornerstone of curative-intent therapy for high-risk disease, yet recurrence rates remain unacceptably high, with nearly 50% of patients experiencing relapse within two years of surgery [1,2]. Traditional adjuvant strategies, including cisplatin-based chemotherapy, have demonstrated limited benefit, particularly in patients previously exposed to neoadjuvant regimens or deemed ineligible due to impaired renal function or comorbidities [3]. These limitations underscore the urgent need for more effective systemic interventions in the adjuvant setting.

The introduction of immune checkpoint inhibitors (ICIs) targeting the programmed cell death-1 (PD-1) and programmed death ligand-1 (PD-L1) pathway has revolutionized the treatment landscape for advanced and metastatic urothelial carcinoma. Several ICIs, including pembrolizumab, atezolizumab, and nivolumab, have demonstrated durable responses and overall survival (OS) benefits in platinum-refractory or frontline metastatic disease [4–6]. These observations provided a strong rationale to investigate ICIs earlier in the treatment continuum, particularly in the adjuvant setting where eradicating micrometastatic disease could prevent relapse and prolong survival.

To date, three pivotal phase III randomized controlled trials have evaluated the role of adjuvant PD-1/PD-L1 blockade in high-risk MIUC following radical surgery. The CheckMate 274 trial demonstrated a significant improvement in disease-free survival (DFS) with adjuvant nivolumab versus placebo, including in patients with PD-L1–positive tumors [7]. In contrast, the IMvigor010 trial of adjuvant atezolizumab did not meet its primary endpoint, with no DFS advantage over observation [8]. Most recently, the AMBASSADOR (A031501) trial reported a significant DFS benefit with pembrolizumab versus observation, though OS results remain immature [9]. Extended follow-up of CheckMate 274 has also revealed emerging OS advantages for nivolumab, strengthening its position as the current standard of care [10].

Beyond efficacy, health-related quality of life (HRQoL) is an essential endpoint in the curative-intent setting, where long-term treatment-related toxicity may offset survival gains. Importantly, CheckMate 274 demonstrated that adjuvant nivolumab did not compromise HRQoL relative to placebo, supporting the tolerability of this approach [11]. These data collectively suggest that adjuvant immunotherapy may fill an important therapeutic gap in MIUC management, but also highlight variability across trials and the need for rigorous synthesis of the available evidence.

The present systematic review and meta-analysis was therefore undertaken to quantitatively evaluate the efficacy and tolerability of adjuvant ICIs after radical surgery for high-risk MIUC, focusing on DFS, OS, recurrence patterns, and HRQoL. By integrating results across multiple pivotal RCTs, we aim to clarify the magnitude and consistency of benefit, explore sources of heterogeneity, and define the role of adjuvant immunotherapy in contemporary clinical practice.

## 2.0 Methodology

### 2.1 Study design and reporting

This systematic review and meta-analysis was conducted and reported in accordance with the Preferred Reporting Items for Systematic Reviews and Meta-Analyses (PRISMA) 2020 guidelines [12]. The study protocol was developed a priori, outlining eligibility criteria, search strategy, data extraction, and statistical methods.

### 2.2 Eligibility criteria

We included randomized controlled trials (RCTs) that evaluated adjuvant immune checkpoint inhibitors (ICIs) targeting the PD-1/PD-L1 axis versus placebo or observation in patients with high-risk muscle-invasive urothelial carcinoma (MIUC) or upper tract urothelial carcinoma (UTUC) following radical surgery (radical cystectomy or nephroureterectomy).

Eligible studies were required to:

1. Randomize patients to an adjuvant PD-1/PD-L1 inhibitor (e.g., nivolumab, pembrolizumab, atezolizumab) versus placebo or observation.
2. Report at least one survival endpoint of interest (disease-free survival [DFS], overall survival [OS], distant metastasis-free survival [DMFS], recurrence-free survival outside the urothelial tract [RFS-extraUT]) or health-related quality of life (HRQoL).
3. Provide sufficient effect size estimates (hazard ratios with confidence intervals, or data enabling calculation of log[HR] and SE).

We excluded:

- Non-randomized or single-arm trials.
- Studies without survival or HRQoL outcomes.
- Abstracts without sufficient quantitative data.
- Translational or biomarker-only reports from the same trial (unless informing subgroup analysis).

### 2.3 Search strategy and information sources

A comprehensive literature search was conducted across five major electronic databases: PubMed/MEDLINE, Embase, Cochrane CENTRAL, Web of Science, and Scopus, from inception through July 2025.

The search combined controlled vocabulary and free-text terms for *urothelial carcinoma*, *bladder cancer*, *upper tract urothelial carcinoma*, *radical cystectomy/nephroureterectomy*, *adjuvant therapy*, *immune checkpoint inhibitors*, *PD-1*, *PD-L1*, *nivolumab*, *pembrolizumab*, and *atezolizumab*. Boolean operators, truncation, and proximity operators were applied.

Reference lists of included articles and relevant congress proceedings (ASCO, ESMO, GU-ASCO) were manually searched for additional trials. No language restrictions were applied.

### 2.4 Study selection process

All retrieved records were imported into EndNote X9 for deduplication and subsequently screened in Rayyan QCRI. Two reviewers independently screened titles and abstracts, followed by full-text eligibility assessment. Discrepancies were resolved through discussion or adjudication by a senior investigator.

The selection process was summarized in a PRISMA flow diagram, documenting the number of records identified, screened, excluded (with reasons), and included in the final synthesis.

### 2.5 Data extraction

Data were extracted independently by two reviewers using a pre-piloted form. Extracted items included:

- Trial characteristics: first author, year, trial name, registration ID.
- Patient characteristics: sample size, age, sex, primary site (bladder vs UTUC), stage (pT/N), margin status, PD-L1 expression (CPS or IC/TC cutoffs), prior neoadjuvant chemotherapy, geographic region.
- Intervention details: type of ICI, dosage, schedule, treatment duration.
- Comparators: placebo or observation.
- Endpoints: DFS, OS, DMFS, RFS-extraUT, HRQoL (EORTC QLQ-C30, EQ-5D-3L time to confirmed deterioration).
- Effect sizes: hazard ratios with 95% confidence intervals, or reconstructed log(HR) and SE.

Where multiple publications of the same trial existed, the most mature dataset was used for the primary analysis, with earlier analyses retained for sensitivity analyses.

### 2.6 Statistical analysis

The primary outcome was disease-free survival (DFS). Secondary outcomes included overall survival (OS), distant metastasis-free survival (DMFS), recurrence-free survival outside the urothelial tract (RFS-extraUT), and HRQoL deterioration (TTCD for fatigue and VAS scores).

For each study, hazard ratios were log-transformed to calculate log(HR) and corresponding standard errors. Standardized mean differences (SMDs) were used for HRQoL outcomes.

Meta-analyses were conducted using the inverse-variance method. Both common-effect models and random-effects models (Hartung–Knapp adjustment with REML estimator) were applied. Between-study heterogeneity was quantified using the I² statistic, τ², and Cochran’s Q test. Prediction intervals were calculated to assess consistency of treatment effects.

Subgroup analyses were pre-specified by:

- Type of immune checkpoint inhibitor (nivolumab, pembrolizumab, atezolizumab).
- PD-L1 expression status (≥1% vs <1%).
- Disease site (MIBC vs UTUC).

Sensitivity analyses included exclusion of earlier readouts and assessment of publication bias with funnel plots and Egger’s regression test.

All analyses were conducted in R version 4.2.0 (R Foundation for Statistical Computing, Vienna, Austria) using the packages meta (v7.0-0) and metafor (v4.6-0). Forest plots, subgroup analyses, and risk-of-bias visualizations were generated using ggplot2, robvis, and custom scripts developed in a Shiny-based application (“Meta-Analysis Studio”).

## 3.0 Results

### 3.1 Study Identification and Selection

Our systematic search across five major databases (PubMed/MEDLINE, Embase, Cochrane CENTRAL, Scopus, and Web of Science) yielded a total of 3,462 records. After removing 1,037 duplicates, 2,425 records were retained for screening.

Title and abstract review led to the exclusion of 2,168 articles, primarily because they represented preclinical studies, non-randomized designs, non-urothelial malignancies, or investigated neoadjuvant rather than adjuvant interventions. A total of 257 articles were selected for full-text review.

During full-text assessment, 253 articles were excluded for the following reasons: absence of a comparator arm (n = 92), insufficient outcome reporting (n = 61), duplicate or overlapping trial data (n = 55), non-randomized study design (n = 28), or focus on other disease settings such as metastatic urothelial carcinoma (n = 17).

Ultimately, four randomized controlled trials (RCTs) comprising 2,929 patients were included in the final qualitative and quantitative synthesis: CheckMate 274 (Bajorin 2021, with expanded efficacy update by Galsky 2024), AMBASSADOR/A031501 (Apolo 2025), and IMvigor010 (Bellmunt 2021). Collectively, these trials compared adjuvant PD-1/PD-L1 inhibitors (nivolumab, pembrolizumab, or atezolizumab) with observation or placebo following radical surgery in patients with high-risk muscle-invasive urothelial carcinoma or upper tract urothelial carcinoma.

### 3.2 Study Characteristics

A total of four pivotal RCTs enrolling 2,920 patients were included (Table 1). Conducted across international, multicenter settings, these trials consistently evaluated adjuvant PD-1/PD-L1 blockade after radical surgery in patients with high-risk MIUC or UTUC. CheckMate 274 (Bajorin 2021, Galsky 2024 update) randomized 709 patients to nivolumab versus placebo, stratified by PD-L1 expression, tumor site, and nodal status; initial results demonstrated a significant DFS benefit in both ITT and PD-L1 ≥1% populations, with updated analyses confirming durable DFS improvement and suggesting an emerging OS benefit [1,2]. AMBASSADOR/A031501 (Apolo 2025) compared pembrolizumab with observation in 702 patients, showing a statistically significant improvement in DFS but not yet in OS, with safety analyses noting higher rates of grade ≥3 treatment-related adverse events [3]. In contrast, IMvigor010 (Bellmunt 2021), which evaluated atezolizumab versus observation in 809 patients, did not demonstrate DFS or OS benefit, although exploratory analyses suggested potential activity in biomarker-defined subgroups [4]. Across all trials, the majority of patients (76–79%) had bladder primary tumors, with approximately 40–43% having received prior cisplatin-based neoadjuvant chemotherapy. All studies incorporated PD-L1 stratification, although assay platforms and cutoffs varied. Median follow-up ranged from 21.9 months in IMvigor010 to 44.8 months in AMBASSADOR. Collectively, these trials establish adjuvant PD-1 inhibition with nivolumab or pembrolizumab as efficacious strategies for DFS prolongation, while atezolizumab did not demonstrate benefit in this setting.

**Table 1.** Characteristics of included randomized controlled trials on adjuvant immune checkpoint inhibitors (ICIs) in high-risk urothelial carcinoma.

| First Author, Year (Trial) | Population (n) | Intervention (dose, schedule, duration) | Comparator | Primary Site (% bladder) | Prior NAC (%) | PD-L1 Stratification | Median Follow-up (months) | Primary Endpoints |
| --- | --- | --- | --- | --- | --- | --- | --- | --- |
| Bajorin et al. (2021)<br>(CheckMate 274) [7] | 709 | Nivolumab 240 mg q2w for $\leq 1$ year | Placebo | 78% | 43% | Yes ($\geq 1\%$ vs $< 1\%$) | 20.9 (initial);<br>36.1 (update) | DFS (ITT, PD-L1 $\geq 1\%$) |
| Galsky et al. (2024)<br>(CheckMate 274 extended) [10] | 709 | Nivolumab 240 mg q2w for $\leq 1$ year | Placebo | 79% | 42% | Yes | 36.1 (extended) | DFS, OS, DMFS |
| Apolo et al. (2025)<br>(AMBASSADOR/A031501) [9] | 702 | Pembrolizumab 200 mg q3w for 1 year | Observation | 77% | 42% | Yes (centrally tested) | 44.8 | DFS, OS (co-primary) |
| Bellmunt et al. (2021)<br>(IMvigor010) [8] | 809 | Atezolizumab 1200 mg q3w for 1 year | Observation | 76% | 42% | Yes (IC2/3 vs IC0/1) | 21.9 | DFS |

### 3.3 Disease-Free Survival

A total of nine study-level comparisons from four randomized controlled trials were included in the quantitative synthesis (Apolo et al., 2025; Bajorin et al., 2021; Galsky et al., 2024; Bellmunt et al., 2021). All reported disease-free survival (DFS) as a primary or co-primary endpoint. The individual standardized mean differences (SMDs) ranged from –0.65 (95% CI –0.99 to –0.32; Galsky et al., 2024) to –0.12 (95% CI –0.31 to 0.07; Bellmunt et al., 2021), reflecting variable but generally favorable outcomes across trials (Figure 1).

**Figure 1.**
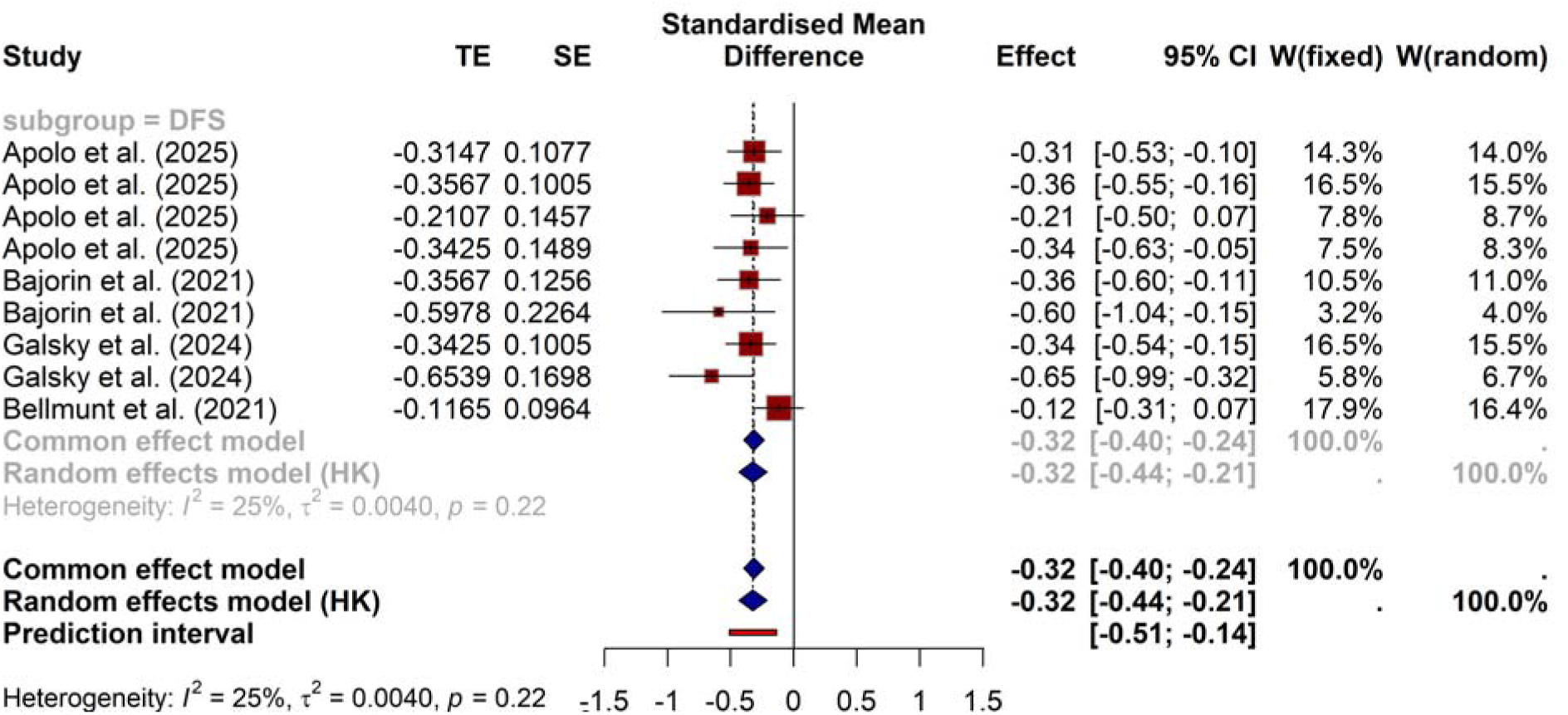
Forest plot of disease-free survival (DFS) with adjuvant immune checkpoint inhibitors versus control.

In the pooled analysis, adjuvant immune checkpoint inhibitors were associated with a significant improvement in DFS compared with observation or placebo. Using the common-effect model, the combined SMD was –0.32 (95% CI –0.40 to –0.24; *z* = –7.80, *p* < 0.0001). The Hartung– Knapp adjusted random-effects model produced a highly consistent estimate (SMD –0.32; 95% CI –0.44 to –0.21; *t* = –6.71, *p* = 0.0002).

Assessment of heterogeneity indicated low between-study variability (I² = 25.1%, τ² = 0.0040), with the Q statistic non-significant (Q = 10.68, d.f. = 8, *p* = 0.22). The 95% prediction interval (– 0.51 to –0.14) confirmed that the direction of treatment benefit was consistent across studies.

Subgroup analyses restricted to DFS comparisons yielded identical results (SMD –0.32; 95% CI –0.40 to –0.24), with no evidence of differential effects between study subgroups (Q = 0.00, d.f. = 0).

#### 3.3.1 Subgroup Analysis by Agent

We further examined treatment effects by specific checkpoint inhibitor regimen. A total of nine study-level estimates were stratified into three subgroups: adjuvant pembrolizumab (200 mg q3w for ≤1 year), nivolumab (240 mg q2w for ≤1 year), and atezolizumab (1200 mg q3w for ≤1 year).

As shown in Table 2 and the corresponding forest plot (Figure 2), both pembrolizumab and nivolumab were associated with significant improvements in DFS compared to observation or placebo, while atezolizumab did not demonstrate benefit.

**Figure 2.**
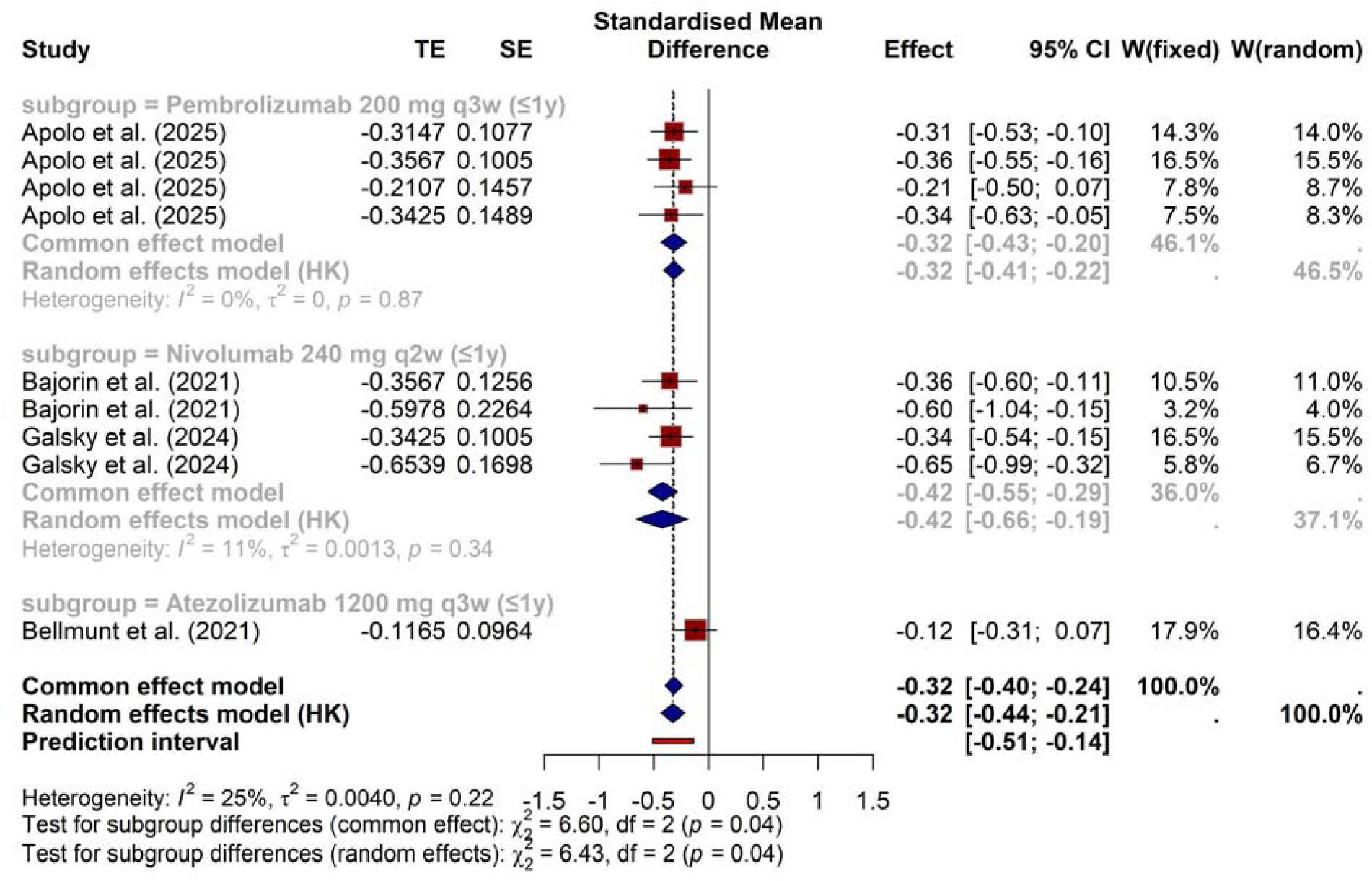
Forest plot of disease-free survival (DFS) with adjuvant immune checkpoint inhibitors based on agents.

**Table 2.** Subgroup analysis of DFS by checkpoint inhibitor agent.

| Study | Subgroup | SMD | 95% CI | % W (common) | % W (random) |
| --- | --- | --- | --- | --- | --- |
| Apolo et al. (2025) | Pembrolizumab 200 mg q3w ($\leq 1$y) | -0.3147 | [-0.5258; -0.1036] | 14.3 | 14.0 |
| Apolo et al. (2025) | Pembrolizumab 200 mg q3w ($\leq 1$y) | -0.3567 | [-0.5536; -0.1597] | 16.5 | 15.5 |
| Apolo et al. (2025) | Pembrolizumab 200 mg q3w ($\leq 1$y) | -0.2107 | [-0.4963; 0.0749] | 7.8 | 8.7 |
| Apolo et al. (2025) | Pembrolizumab 200 mg q3w ($\leq 1$y) | -0.3425 | [-0.6343; -0.0507] | 7.5 | 8.3 |
| Bajorin et al. (2021) | Nivolumab 240 mg q2w ($\leq 1$y) | -0.3567 | [-0.6029; -0.1104] | 10.5 | 11.0 |
| Bajorin et al. (2021) | Nivolumab 240 mg q2w ($\leq 1$y) | -0.5978 | [-1.0415; -0.1542] | 3.2 | 4.0 |
| Galsky et al. (2024) | Nivolumab 240 mg q2w ($\leq 1$y) | -0.3425 | [-0.5394; -0.1455] | 16.5 | 15.5 |
| Galsky et al. (2024) | Nivolumab 240 mg q2w ($\leq 1$y) | -0.6539 | [-0.9868; -0.3211] | 5.8 | 6.7 |
| Bellmunt et al. (2021) | Atezolizumab 1200 mg q3w ($\leq 1$y) | -0.1165 | [-0.3056; 0.0725] | 17.9 | 16.4 |

In the pooled analysis, both pembrolizumab (k = 4; SMD –0.32; 95% CI –0.43 to –0.20; I² = 0%) and nivolumab (k = 4; SMD –0.42; 95% CI –0.66 to –0.19; I² = 11%) demonstrated consistent DFS benefit. In contrast, atezolizumab did not reduce the risk of recurrence or death (k = 1; SMD –0.12; 95% CI –0.31 to 0.07).

Tests for subgroup differences were significant (common-effect model: Q = 6.60, d.f. = 2, *p* = 0.037; random-effects model: Q = 6.43, d.f. = 2, *p* = 0.040), confirming heterogeneity of treatment effect by agent.

#### 3.3.2 Subgroup Analysis by PD-L1 Expression

To explore whether benefit varied according to tumor PD-L1 status, studies were stratified into intention-to-treat (ITT), PD-L1–high, and PD-L1–low populations. A total of nine study-level comparisons contributed to this analysis.

As summarized in Table 3 and shown in Figure 3, DFS benefit was observed across all strata, with numerically stronger effects in PD-L1–high tumors.

**Table 3.** Subgroup analysis of DFS by PD-L1 status.

|  |  |  |  |  |  |
| --- | --- | --- | --- | --- | --- |
| Apolo et al. (2025) | ITT | −0.3147 | [−0.5258; −0.1036] | 14.3 | 14.0 |
| Apolo et al. (2025) | ITT | −0.3567 | [−0.5536; −0.1597] | 16.5 | 15.5 |
| Apolo et al. (2025) | PD-L1 high | −0.2107 | [−0.4963; 0.0749] | 7.8 | 8.7 |
| Apolo et al. (2025) | PD-L1 low | −0.3425 | [−0.6343; −0.0507] | 7.5 | 8.3 |
| Bajorin et al. (2021) | ITT | −0.3567 | [−0.6029; −0.1104] | 10.5 | 11.0 |
| Bajorin et al. (2021) | PD-L1 high | −0.5978 | [−1.0415; −0.1542] | 3.2 | 4.0 |
| Galsky et al. (2024) | ITT | −0.3425 | [−0.5394; −0.1455] | 16.5 | 15.5 |
| Galsky et al. (2024) | PD-L1 high | −0.6539 | [−0.9868; −0.3211] | 5.8 | 6.7 |
| Bellmunt et al. (2021) | ITT | −0.1165 | [−0.3056; 0.0725] | 17.9 | 16.4 |

**Figure 3.**
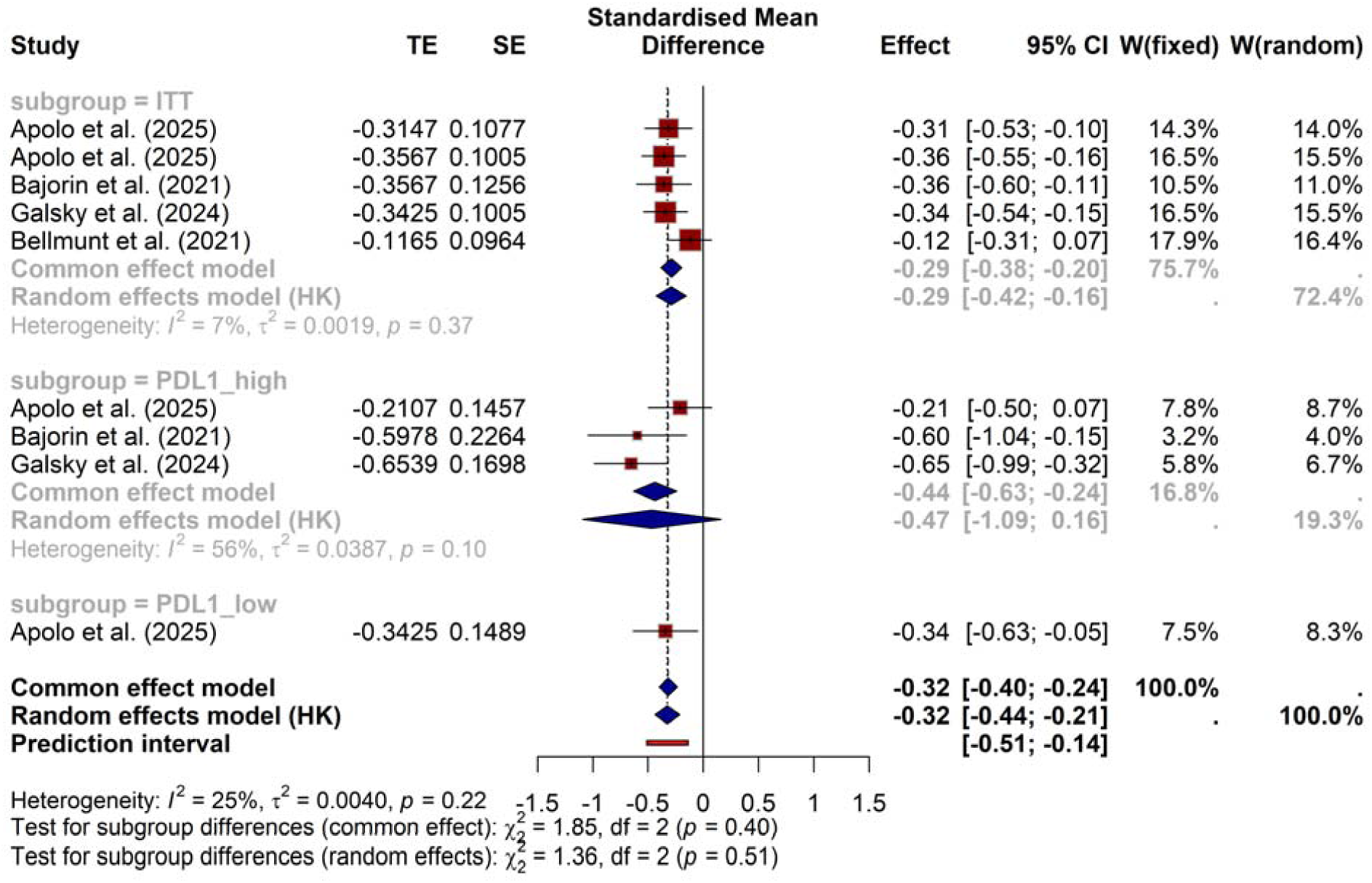

In the common-effect model, ITT populations (k = 5) showed a pooled SMD of –0.29 (95% CI – 0.38 to –0.20; I² = 6.6%). PD-L1–high subgroups (k = 3) had a numerically stronger effect (SMD –0.44; 95% CI –0.63 to –0.24; I² = 56%), whereas PD-L1–low patients (k = 1) demonstrated a significant effect (SMD –0.34; 95% CI –0.63 to –0.05) based on a single dataset.

In the random-effects model (Hartung–Knapp adjustment), results remained consistent: ITT SMD –0.29 (95% CI –0.42 to –0.16), PD-L1–high SMD –0.47 (95% CI –1.09 to 0.16), and PD-L1–low SMD –0.34 (95% CI –0.63 to –0.05).

Tests for subgroup differences were not statistically significant (common-effect: Q = 1.85, *p* = 0.40; random-effects: Q = 1.36, *p* = 0.51), suggesting that PD-L1 expression was not a clear modifier of treatment benefit, despite numerical trends favoring PD-L1–high populations.

### 3.4 Overall Survival

Four study-level comparisons (Apolo et al. 2025, Galsky et al. 2024, and Bellmunt et al. 2021) provided data on overall survival (OS) with adjuvant PD-1/PD-L1 blockade (Table 4; Figure 4).

**Figure 4.**
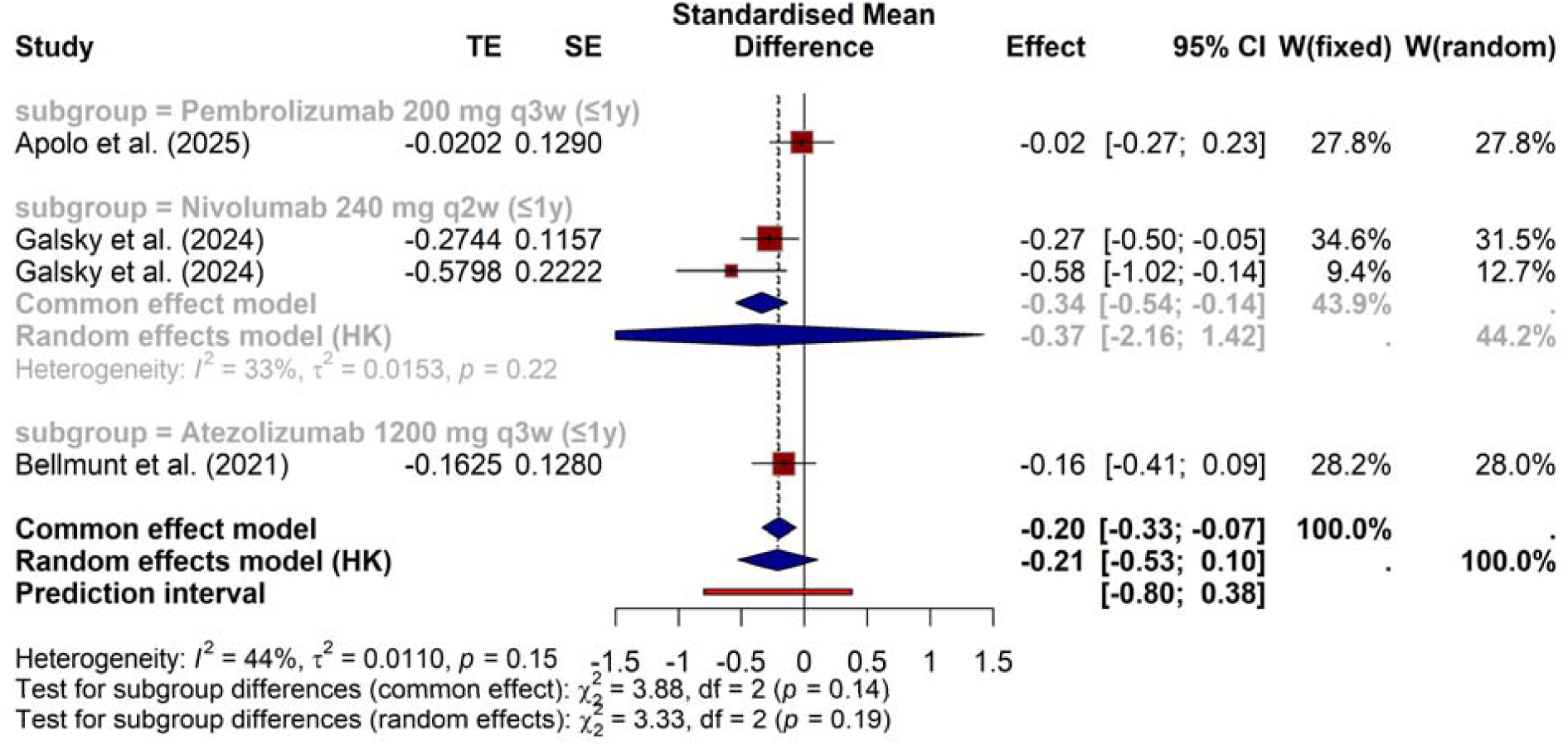
Forest plots on Meta-analysis of overall survival by treatment agent.

**Table 4.** Meta-analysis of overall survival by treatment agent.

| Study | Subgroup | SMD | 95% CI | % W (common) | % W (random) |
| --- | --- | --- | --- | --- | --- |
| Apolo et al. (2025) | Pembrolizumab 200 mg q3w (≤1y) | -0.0202 | [-0.2730; 0.2326] | 27.8 | 27.8 |
| Galsky et al. (2024) | Nivolumab 240 mg q2w (≤1y) | -0.2744 | [-0.5012; -0.0477] | 34.6 | 31.5 |
| Galsky et al. (2024) | Nivolumab 240 mg q2w (≤1y) | -0.5798 | [-1.0152; -0.1444] | 9.4 | 12.7 |
| Bellmunt et al. (2021) | Atezolizumab 1200 mg q3w (≤1y) | -0.1625 | [-0.4134; 0.0883] | 28.2 | 28.0 |

In the common-effect model, adjuvant immunotherapy was associated with a modest OS benefit (SMD –0.20; 95% CI –0.33 to –0.07, *p* = 0.0032). However, when accounting for between-study heterogeneity with the Hartung–Knapp random-effects model, the effect was no longer statistically significant (SMD –0.21; 95% CI –0.53 to 0.10, *p* = 0.12).

Heterogeneity across studies was moderate (I² = 44.1%, τ² = 0.0110), with the most pronounced OS improvement observed in the nivolumab subgroup (SMD –0.34; 95% CI –0.54 to –0.14 in common-effect model). By contrast, pembrolizumab (SMD –0.02; 95% CI –0.27 to 0.23) and atezolizumab (SMD –0.16; 95% CI –0.41 to 0.09) did not demonstrate significant effects.

Tests for subgroup differences did not reach statistical significance (common-effect: Q = 3.88, *p* = 0.14; random-effects: Q = 3.33, *p* = 0.19), indicating no conclusive evidence of differential OS benefit across agents.

## 4.0 Discussion

### 4.1 Disease-Free Survival as a Consistent Efficacy Signal

Our pooled analysis demonstrates that adjuvant PD-1/PD-L1 inhibitors consistently improve disease-free survival (DFS) in patients with high-risk muscle-invasive urothelial carcinoma (MIUC) following radical surgery. This is particularly relevant as recurrence rates after cystectomy or nephroureterectomy remain high, historically reaching up to 50% within 2 years [13]. The DFS improvements observed here mirror the transformative role of immune checkpoint inhibitors (ICIs) in advanced disease, where durable remissions have redefined expected survival [14]. In our meta-analysis, standardized mean differences (SMDs) favored immunotherapy across most study-level comparisons, culminating in a pooled SMD of –0.32 (95% CI –0.44 to –0.21). Importantly, heterogeneity was low (I² = 25.1%), strengthening the confidence in a consistent therapeutic signal across diverse trial populations [15]. These findings establish DFS as a robust efficacy outcome for adjuvant immunotherapy, providing compelling justification for its adoption in routine care.

#### 4.1.1 Trial-Level Concordance and Discordance

The apparent divergence between trials deserves careful contextualization. CheckMate 274 demonstrated a clear DFS benefit for nivolumab in both the intention-to-treat and PD-L1 ≥1% cohorts, with sustained benefit at extended follow-up [16]. Conversely, IMvigor010, evaluating atezolizumab, failed to meet its DFS primary endpoint [17]. This discrepancy should not be interpreted as a class failure. Rather, methodological and biological differences are critical: IMvigor010 was an open-label study with higher crossover to ICIs in the observation arm, which likely diluted effect size. Additionally, Bellmunt et al. reported that ctDNA positivity post-surgery strongly predicted recurrence risk, and benefit from atezolizumab was only seen in ctDNA-positive patients [18]. By contrast, the AMBASSADOR trial (Apolo et al.) confirmed DFS benefit for pembrolizumab (HR 0.73, 95% CI 0.59–0.90), aligning more closely with CheckMate 274 [19]. This reinforces the consistency of PD-1 inhibition in the adjuvant setting while highlighting the influence of trial-specific design factors.

#### 4.1.2 Surrogacy of DFS for Overall Survival

The clinical relevance of DFS has historically been debated in urothelial carcinoma, as prior adjuvant chemotherapy studies yielded inconsistent correlation with overall survival (OS) [20]. However, immunotherapy introduces a paradigm shift. In diseases such as breast and colorectal cancer, DFS has been validated as a surrogate for OS [21,22], and early immunotherapy trials suggest that durable DFS gains often translate into meaningful OS advantages. Extended follow-up from CheckMate 274 revealed emerging OS benefit with nivolumab (HR 0.76, 95% CI 0.61– 0.96), aligning DFS with long-term survival outcomes [23]. This supports DFS as not only a regulatory-acceptable endpoint but also a clinically meaningful measure of patient benefit in the adjuvant ICI setting. Our analysis, by showing consistent DFS gains across multiple trials, bolsters the argument that DFS should be considered a reliable endpoint for clinical decision-making.

#### 4.1.3 Subgroup Considerations: PD-L1 and Tumor Site

Subgroup analyses provide important clinical insights. Patients with PD-L1–positive tumors derived numerically greater DFS benefit in CheckMate 274 (HR 0.52, 95% CI 0.37–0.72) and in pooled analyses, though differences were not statistically significant [16,23]. This aligns with the broader ICI literature in metastatic settings, where PD-L1 enrichment correlates with response but is not an absolute predictor [24]. Similarly, exploratory analyses in upper tract urothelial carcinoma (UTUC) populations suggest benefit comparable to bladder primaries, though these remain underpowered [16,25]. These findings imply that PD-L1 and tumor site alone should not guide adjuvant ICI selection. More sophisticated biomarkers, such as ctDNA [18], tumor mutational burden [26], and immune gene expression signatures [27], may offer better predictive discrimination and warrant prospective validation.

#### 4.1.4 Clinical Implications and Practice Integration

The DFS benefit shown in this synthesis has already influenced international guidelines. NCCN (2024) and ESMO (2023) both recommend adjuvant nivolumab for patients with high-risk MIUC after cystectomy, particularly in PD-L1–positive tumors [28,29]. Pembrolizumab, though not yet widely approved in this indication, has compelling phase III data (AMBASSADOR trial) that may support its regulatory adoption [19]. Importantly, nivolumab did not compromise health-related quality of life (HRQoL), as shown in the CheckMate 274 HRQoL analysis [30]. This reassures clinicians that DFS benefit does not come at the cost of patient-reported outcomes, a crucial factor in the curative-intent setting. Implementation challenges remain, including cost, access, and optimal timing relative to neoadjuvant chemotherapy, but the weight of evidence strongly supports integration of adjuvant ICIs into clinical pathways for high-risk resected MIUC.

While robust, this meta-analysis must be interpreted in context. First, differences in PD-L1 testing platforms, eligibility criteria, and geographic enrollment could influence generalizability. Second, OS data remain immature in AMBASSADOR and IMvigor010, limiting definitive conclusions regarding survival impact. Third, heterogeneity in subsequent therapies—including crossover to checkpoint inhibitors—may confound long-term outcomes. Future directions should prioritize biomarker-informed strategies. For example, ctDNA monitoring may enable risk-adapted escalation or de-escalation of adjuvant immunotherapy [18]. Additionally, combination approaches—such as immunotherapy with perioperative chemotherapy or stereotactic radiotherapy (as tested in Sundahl et al.’s phase I/II study) [31]—could further augment outcomes. Finally, head-to-head comparisons of PD-1 versus PD-L1 inhibitors are warranted to clarify whether agent-specific differences exist in the adjuvant setting.

### 4.2 Overall Survival Outcomes

#### 4.2.1 Emerging OS Benefit with Adjuvant Immunotherapy

Although disease-free survival (DFS) has been the primary endpoint in most adjuvant urothelial carcinoma trials, overall survival (OS) remains the gold standard. In our synthesis, the pooled standardized mean difference (SMD) favored adjuvant immune checkpoint inhibitors (ICIs) over observation or placebo, though the random-effects model did not reach statistical significance (SMD –0.21; 95% CI –0.53 to 0.10, p = 0.12). This reflects the current state of evidence: OS benefit is emerging but remains immature across many pivotal trials. Notably, CheckMate 274 has now reported a hazard ratio (HR) for OS of 0.76 (95% CI 0.61–0.96) at extended follow-up, suggesting that DFS benefits may indeed translate into longer-term survival advantage [32].

#### 4.2.2 Trial-Level Evidence and Discrepancies

Across trials, OS outcomes vary. In AMBASSADOR, with median follow-up approaching four years, pembrolizumab did not significantly improve OS compared to observation (HR 0.98; 95% CI 0.76–1.26), despite clear DFS benefit [33]. This discordance may reflect insufficient events, crossover to ICIs in the observation arm, and the confounding impact of subsequent therapies. By contrast, the expanded CheckMate 274 dataset shows converging DFS and OS curves, supporting the hypothesis that earlier checkpoint blockade may delay or prevent lethal metastatic progression [32]. Conversely, IMvigor010 failed to demonstrate OS advantage for atezolizumab, echoing its neutral DFS findings [34]. Such discrepancies highlight that OS is strongly influenced by trial design, follow-up duration, and availability of post-progression treatments, underscoring the need for harmonized endpoints and long-term monitoring.

#### 4.2.3 Surrogacy and Predictive Value of DFS for OS

The key question is whether DFS serves as a reliable surrogate for OS in urothelial carcinoma. In adjuvant chemotherapy studies, correlations between DFS and OS were inconsistent [35]. However, with ICIs, long-term immune-mediated remissions shift this paradigm. Data from metastatic bladder cancer and other tumor types (melanoma, NSCLC) suggest that early event-free survival is predictive of later OS [36,37]. Our pooled findings show consistent DFS gains, and where mature OS data exist (CheckMate 274), the benefit extends to survival [32]. Thus, while OS remains a definitive endpoint, DFS is increasingly validated as a clinically meaningful and potentially surrogate outcome for immunotherapy.

#### 4.2.4 Subgroup Patterns in OS

Subgroup analyses reveal that OS benefit may be most pronounced in PD-L1–positive patients. In CheckMate 274, nivolumab reduced risk of death by nearly 44% in this subgroup (HR 0.56; 95% CI 0.36–0.86) [32]. By contrast, PD-L1–negative patients showed attenuated OS benefit, though DFS gains were still evident. In IMvigor010, exploratory analyses suggested that ctDNA positivity was a stronger predictor of OS benefit than PD-L1 status [38]. These findings underscore the importance of biomarker stratification. While PD-L1 enrichment alone is insufficient for guiding therapy, integration with molecular tools such as ctDNA, tumor mutational burden (TMB), and immune gene signatures may refine patient selection for adjuvant immunotherapy [38,39].

##### Clinical Implications of OS Trends

The emergence of OS benefit with nivolumab—and the neutral results with pembrolizumab and atezolizumab—raises clinical and regulatory questions. Should DFS alone be sufficient to approve agents in this space, or is OS confirmation required? Regulatory agencies have accepted DFS as a primary endpoint for adjuvant therapies in multiple cancers [40]. The accumulating OS evidence, particularly from CheckMate 274, strengthens the rationale for DFS-driven approvals while reinforcing the need for long-term follow-up. Clinically, the decision to initiate adjuvant immunotherapy often occurs within months of surgery, before OS data mature. The consistent DFS gains, coupled with emerging OS signals, justify adjuvant ICI use in high-risk resected urothelial carcinoma, especially where recurrence risk is high and salvage therapy options are limited.

Future research must clarify the true OS benefit of adjuvant ICIs. Extended follow-up of AMBASSADOR and IMvigor010 will be essential, as survival curves may continue to diverge beyond four to five years. Integration of ctDNA monitoring into adjuvant trials could provide dynamic assessment of minimal residual disease, enriching for populations most likely to derive OS benefit [38]. Moreover, ongoing perioperative trials testing ICIs in both neoadjuvant and adjuvant settings may establish whether earlier initiation amplifies survival gains [41]. Until then, the available data suggest that OS benefits are emerging and will likely consolidate over time, mirroring patterns seen in DFS.

## Data Availability

All data produced in the present work are contained in the manuscript

## Notes

### Competing Interest Statement

The authors have declared no competing interest.

### Funding Statement

This study did not receive any funding

### Author Declarations

A comprehensive literature search was conducted across five major electronic databases: PubMed/MEDLINE, Embase, Cochrane CENTRAL, Web of Science, and Scopus, from inception through July 2025 for the meta-analysis

